# Metabolomic Biomarkers and Signature of Ultra-processed Foods Intake and Cardiovascular Morbidity and Mortality

**DOI:** 10.1101/2025.08.29.25334742

**Authors:** Lei Wang, Jiaqi Yang, Jingsha Chen, Casey M. Rebholz, Xiao-Ou Shu, Martha J. Shrubsole, Deepak K. Gupta, Loren Lipworth, Siyuan Ma, Qi Dai, Wei Zheng, Danxia Yu

## Abstract

Ultra-processed foods (UPFs) intake may increase cardiovascular disease (CVD) risk, but the mechanisms remain unclear and evidence on UPF biomarkers is limited. Leveraging untargeted blood metabolomics in three prospective cohorts, we identified and validated circulating metabolites associated with UPF intake and CVD risk and mortality. Discovery was conducted in the Southern Community Cohort Study (N=1,688), with validation in the Prostate, Lung, Colorectal, and Ovarian Cancer Screening Trial (N=2,315) and the Atherosclerosis Risk in Communities Study (N=3,682). We identified (n=142) and validated (n=43) metabolites associated with UPF intake, with several of these metabolites further linked to incident coronary heart disease (n=5), CVD mortality (n=2), and total mortality (n=20). Importantly, we developed a metabolite signature for UPF intake, which demonstrated strong associations with outcomes (OR/HR=1.25–1.46 per 1-SD increase) and explained 59–77% of the UPF-disease associations. These findings may enhance the assessment and mechanistic understanding of how UPFs impact human health.

## Introduction

Ultra-processed foods (UPFs) — industrial formulations typically made with little or no whole foods and abundant in additives such as colorings, flavorings, emulsifiers, and preservatives — have become increasingly dominant contributors to the modern diet^1^. In the United States, UPFs account for 50–70% of total daily energy intake^2^. Characterized by high levels of added sugars, saturated fats, and sodium and low levels of fiber and micronutrients, UPFs have raised substantial public health concerns. Accumulating epidemiological evidence links higher UPF consumption with poor metabolic health, including obesity, high waist circumference, and low HDL-cholesterol level^3^, as well as a 29–35% greater cardiovascular disease (CVD) risk and a 21–25% higher total mortality^4,5^.

Observational studies on UPFs and their health effects have faced challenges, however, related to dietary assessment errors and unclear biological pathways by which UPFs may impact health. Hypothesized mechanisms include disruption of the gut microbiome, systemic inflammation, oxidative stress, and metabolic dysregulation triggered by the nutrient-poor composition and synthetic additives in UPF^6^. Large-scale blood metabolomics can provide a more comprehensive picture of the underlying metabolic profile associated with UPF intake and further link them to disease outcomes, particularly CVD, making it a powerful tool for precision nutrition and mechanistic investigation. Numerous circulating metabolites from various chemical classes have been consistently associated with the risk of CVD or total mortality, including amino acids (e.g., branched-chain and aromatic amino acids, tryptophan, and imidazole propionate), lipids (e.g., lysophophatidylcholines, sphingomyelins, and microbiome-derived short-chain fatty acids, secondary bile acids, and trimethylamine N-oxide), and nucleotides (e.g., N2,N2-dimethylguanosine, N6 succinyladenosine, and pseudouridine from purine metabolism)^7–11^. However, the metabolite profile of UPF intake has not been well elucidated. Previous studies have identified 12 to 257 metabolites associated with UPF intake, with substantial variations across findings^12–17^. In addition, none of these studies have further evaluated the associations between UPF-related metabolites and incident CVD.

To address these gaps, we conducted a two-stage study leveraging untargeted blood metabolomics data from three large, well-characterized prospective cohorts in the United States. First, we identified individual blood metabolites related to UPF intake and evaluated the associations of UPF-related metabolites with coronary heart disease (CHD) incidence, CVD mortality, and total mortality in the Southern Community Cohort Study (SCCS, N=1688). Then, we validated the UPF-related metabolites identified above and their associations with disease outcomes in the Prostate, Lung, Colorectal, and Ovarian Cancer Screening Trial (PLCO; N=2,315) and the Atherosclerosis Risk in Communities Study (ARIC; N=3,682). In addition, we constructed a metabolite signature (MetSig)—a score derived from UPF-related metabolites in the SCCS—to capture the overall metabolite profile of UPF intake and evaluate its associations with these outcomes. This study aimed to advance our understanding of the mechanisms underlying UPF intake and human health and improve the precision in evaluating the relations between UPF intake and health outcomes.

## Results

### Participants’ characteristics

The discovery stage included 1,688 participants from two metabolomic nested case-control studies (incident CHD and prostate cancer) within the SCCS. The mean enrollment age of included participants was 55.1 (SD: 7.9) years, with 70% male and 61% self-reported Black/ African American adults (**Table 1**). The mean total UPF intake was 41.1% (SD: 15.3%) of total daily food intake by weight. Participants with higher UPF intake were more likely to be younger, male, Black, current smokers, and alcohol drinkers, and have lower education and income levels, as well as higher daily energy intake (all P<0.05). However, a higher prevalence of hypercholesterolemia was observed among participants with lower UPF intake, suggesting possible dietary changes after disease diagnosis.

**Table 1.**
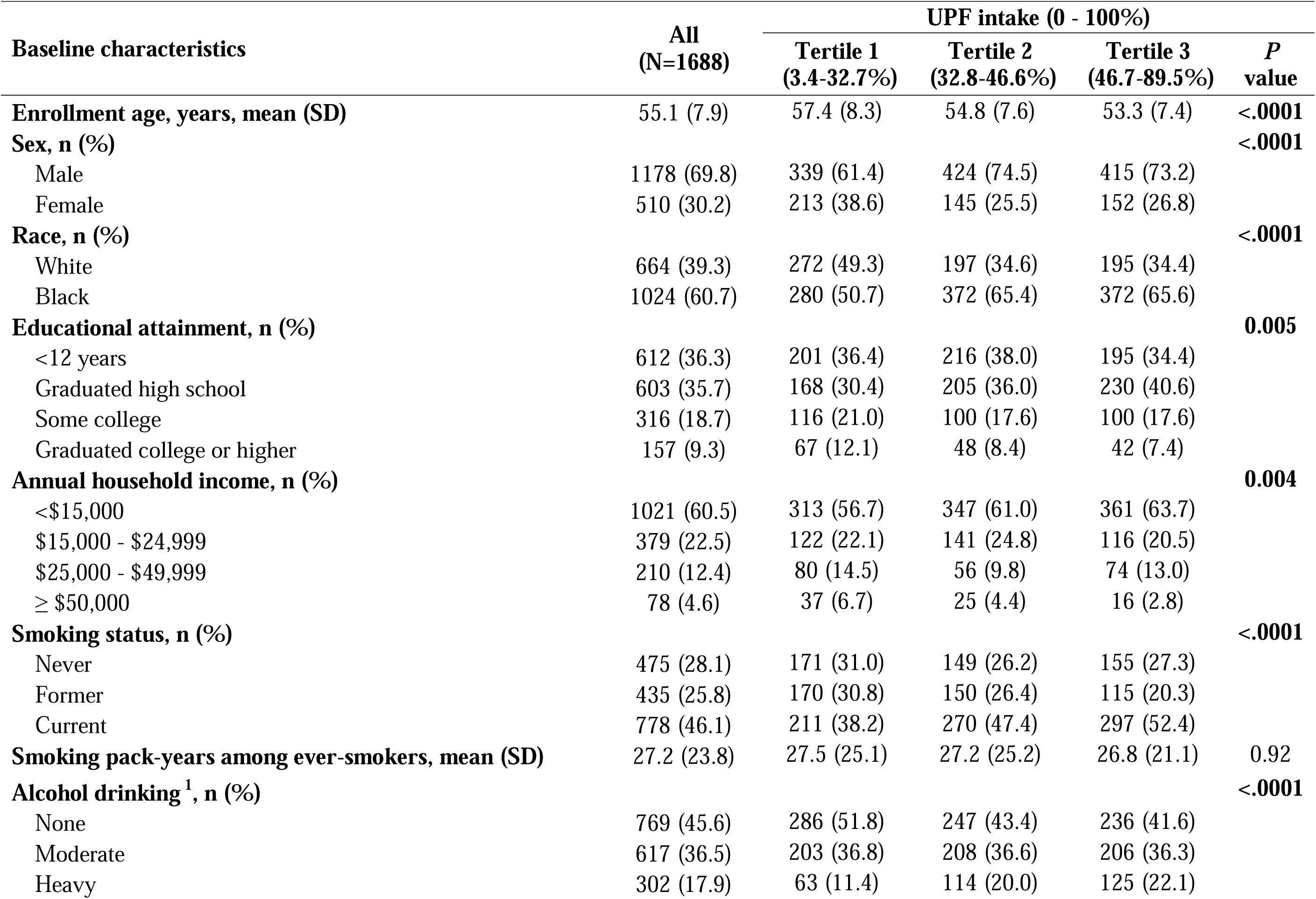
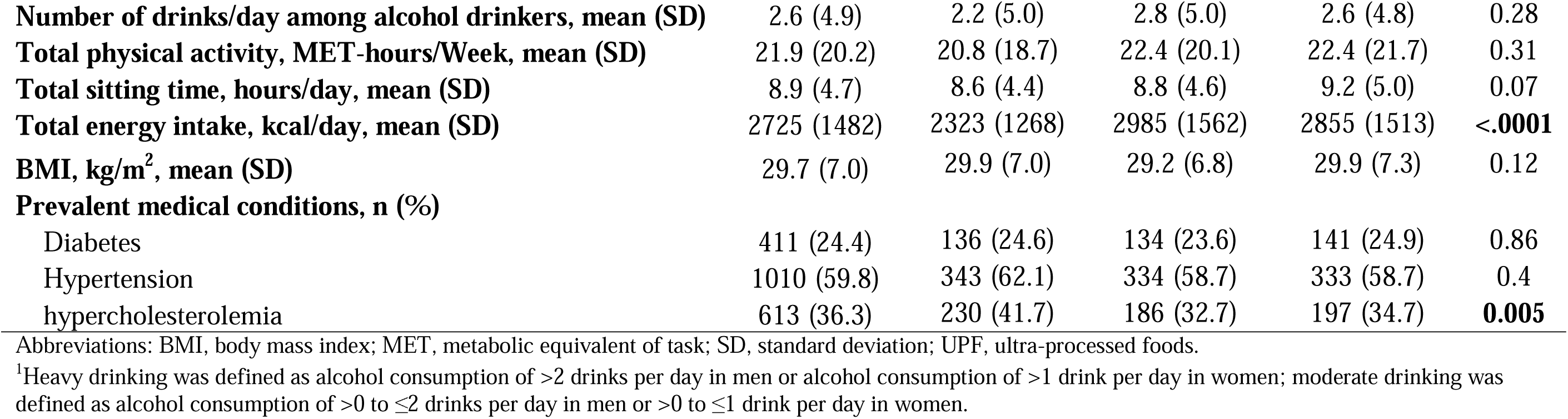
Baseline characteristics of study participants across UPF tertiles in the SCCS.

| Baseline characteristics | All<br>(N=1688) | UPF intake (0 - 100%) |  |  | *P*<br>value |
| --- | --- | --- | --- | --- | --- |
|  |  | Tertile 1<br>(3.4-32.7%) | Tertile 2<br>(32.8-46.6%) | Tertile 3<br>(46.7-89.5%) |  |
| **Enrollment age, years, mean (SD)** | 55.1 (7.9) | 57.4 (8.3) | 54.8 (7.6) | 53.3 (7.4) | **<.0001** |
| **Sex, n (%)** |  |  |  |  | **<.0001** |
| Male | 1178 (69.8) | 339 (61.4) | 424 (74.5) | 415 (73.2) |  |
| Female | 510 (30.2) | 213 (38.6) | 145 (25.5) | 152 (26.8) |  |
| **Race, n (%)** |  |  |  |  | **<.0001** |
| White | 664 (39.3) | 272 (49.3) | 197 (34.6) | 195 (34.4) |  |
| Black | 1024 (60.7) | 280 (50.7) | 372 (65.4) | 372 (65.6) |  |
| **Educational attainment, n (%)** |  |  |  |  | **0.005** |
| <12 years | 612 (36.3) | 201 (36.4) | 216 (38.0) | 195 (34.4) |  |
| Graduated high school | 603 (35.7) | 168 (30.4) | 205 (36.0) | 230 (40.6) |  |
| Some college | 316 (18.7) | 116 (21.0) | 100 (17.6) | 100 (17.6) |  |
| Graduated college or higher | 157 (9.3) | 67 (12.1) | 48 (8.4) | 42 (7.4) |  |
| **Annual household income, n (%)** |  |  |  |  | **0.004** |
| <\$15,000 | 1021 (60.5) | 313 (56.7) | 347 (61.0) | 361 (63.7) | |
| \$15,000 - \$24,999 | 379 (22.5) | 122 (22.1) | 141 (24.8) | 116 (20.5) | |
| \$25,000 - \$49,999 | 210 (12.4) | 80 (14.5) | 56 (9.8) | 74 (13.0) | |
| ≥ \$50,000 | 78 (4.6) | 37 (6.7) | 25 (4.4) | 16 (2.8) | |
| **Smoking status, n (%)** |  |  |  |  | **<.0001** |
| Never | 475 (28.1) | 171 (31.0) | 149 (26.2) | 155 (27.3) |  |
| Former | 435 (25.8) | 170 (30.8) | 150 (26.4) | 115 (20.3) |  |
| Current | 778 (46.1) | 211 (38.2) | 270 (47.4) | 297 (52.4) |  |
| **Smoking pack-years among ever-smokers, mean (SD)** | 27.2 (23.8) | 27.5 (25.1) | 27.2 (25.2) | 26.8 (21.1) | 0.92 |
| **Alcohol drinking<sup>1</sup>, n (%)** |  |  |  |  | **<.0001** |
| None | 769 (45.6) | 286 (51.8) | 247 (43.4) | 236 (41.6) |  |
| Moderate | 617 (36.5) | 203 (36.8) | 208 (36.6) | 206 (36.3) |  |
| Heavy | 302 (17.9) | 63 (11.4) | 114 (20.0) | 125 (22.1) |  |
| **Number of drinks/day among alcohol drinkers, mean (SD)** | 2.6 (4.9) | 2.2 (5.0) | 2.8 (5.0) | 2.6 (4.8) | 0.28 |
| **Total physical activity, MET-hours/Week, mean (SD)** | 21.9 (20.2) | 20.8 (18.7) | 22.4 (20.1) | 22.4 (21.7) | 0.31 |
| **Total sitting time, hours/day, mean (SD)** | 8.9 (4.7) | 8.6 (4.4) | 8.8 (4.6) | 9.2 (5.0) | 0.07 |
| **Total energy intake, kcal/day, mean (SD)** | 2725 (1482) | 2323 (1268) | 2985 (1562) | 2855 (1513) | **<.0001** |
| **BMI, kg/m<sup>2</sup>, mean (SD)** | 29.7 (7.0) | 29.9 (7.0) | 29.2 (6.8) | 29.9 (7.3) | 0.12 |
| **Prevalent medical conditions, n (%)** |  |  |  |  |  |
| Diabetes | 411 (24.4) | 136 (24.6) | 134 (23.6) | 141 (24.9) | 0.86 |
| Hypertension | 1010 (59.8) | 343 (62.1) | 334 (58.7) | 333 (58.7) | 0.4 |
| hypercholesterolemia | 613 (36.3) | 230 (41.7) | 186 (32.7) | 197 (34.7) | **0.005** |

Abbreviations: BMI, body mass index; MET, metabolic equivalent of task; SD, standard deviation; UPF, ultra-processed foods.
<sup>1</sup>Heavy drinking was defined as alcohol consumption of >2 drinks per day in men or alcohol consumption of >1 drink per day in women; moderate drinking was defined as alcohol consumption of >0 to ≤2 drinks per day in men or >0 to ≤1 drink per day in women.

The validation stage included 2,315 participants from four metabolomic case-control studies of cancers nested within the PLCO and 3,682 participants with metabolomics data from the ARIC study. SCCS, PLCO, and ARIC all conducted metabolite profiling in blood samples collected before cancer/CVD diagnosis using the Metabolon untargeted global discovery assay (see **Methods**). PLCO participants had a mean age of 63.0 (SD: 5.1) years, with 44.6% male and 91.5% White adults. ARIC participants had a mean age of 53.4 (SD: 5.7) years, with 39.1% male and 37.0% White adults (**Supplementary Table 1**). The average UPF intake was lower in PLCO compared to SCCS (22.7% vs. 41.1% of total daily food intake by weight); while in ARIC study, the absolute UPF intake was assessed with a mean of 5.6 (SD: 3.0) servings/day. In addition, compared to SCCS participants, PLCO and ARIC participants had higher education levels and lower smoking rates, daily energy intake, and prevalence of metabolic diseases.

### UPF-related circulating metabolites

For discovery, we identified 142 metabolites associated with UPF intake in the SCCS after multivariable adjustment (all *FDR*<0.1; **Fig. 1**). The majority were xenobiotics (n=38) or lipids (n=29), followed by amino acids (n=19), nucleotides (n=7), cofactors and vitamins (n=5), peptides (n=3), carbohydrates (n=3), energy metabolites (n=2), and partially characterized molecules (n=2); 34 were unknown metabolites.

**Fig. 1.**
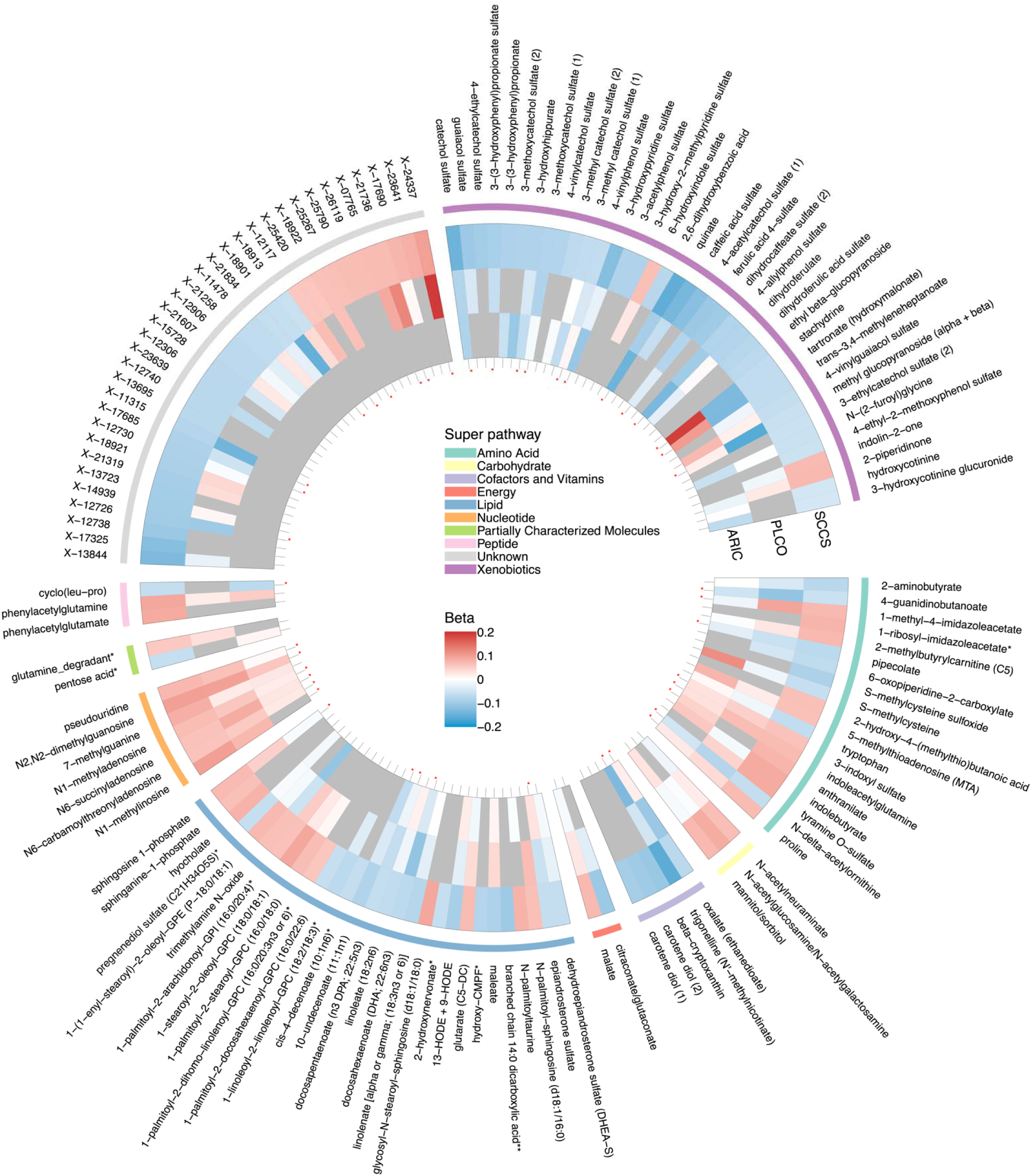
Discovery and Validation of UPF-related metabolites in SCCS, PLCO, and ARIC. Linear regression was used to assess the associations between UPF score (per 1-SD increase) and individual metabolites. Covariates included: 1) for the SCCS — fasting status, batch, enrollment age, sex, race, education, income, smoking status, pack years, number of alcohol drinks per day, physical activity, sitting hours, daily calories intake, BMI, and history of diabetes, hypertension, and hypercholesterolemia at baseline; 2) for the PLCO — study batch, case-control status, enrollment age, sex, race, education, smoking, alcohol drinking status, vigorous activity, daily caloric intake, BMI, and history of diabetes and hypertension at baseline; and 3) for the ARIC study — enrollment age, sex, race, center, education, income, smoking, alcohol consumption, physical activity index, daily caloric intake, BMI, and history of diabetes, hypertension, and lipid-lowering medication use at baseline. Metabolites associated with UPF intake at FDR <0.1 in the SCCS were presented (a total of 142 metabolites). Cells in grey indicated metabolites unavailable in PLCO or ARIC. A red star indicated that the metabolite was validated in PLCO or ARIC.

Among these 142 metabolites, 104 metabolites were available in PLCO or ARIC for validation, and 43 metabolites showed significant associations with UPF intake in the same direction as observed at the discovery stage (P<0.05 in either validation cohort; **Fig. 1**). The validated metabolites were mainly xenobiotics (n=12), amino acids (n=8), nucleotides (n=6), or lipids (n=5), including positively associated metabolites involved in histidine (1-methyl-4-imidazoleacetate), tryptophan (pipecolate, anthranilate, and indoleacetylglutamine), and purine metabolism (e.g., N2,N2-dimethylguanosine, N1-methyladenosine, and N6-succinyladenosine) and negatively associated metabolites from long chain polyunsaturated fatty acids (docosapentaenoate [DPA] and docosahexaenoate [DHA]) and plant-based food/polyphenols (e.g., 4-vinylphenol sulfate, 3-methoxycatechol sulfate, and catechol sulfate).

### UPF-related metabolites and CHD incidence

The 142 UPF-related metabolites identified in the SCCS were further examined for their associations with CHD incidence, CVD mortality, and total mortality, adjusting for sociodemographics, lifestyle behaviors, and metabolic health conditions (see **Methods**); metabolites showing statistically significant associations (i.e., FDR<0.1) were carried forward to the validation stage. In the nested case-control study (511 age-/sex-/race-matched pairs) within the SCCS, 14 metabolites were significantly associated with increased CHD risk (OR=1.19 to 1.41 per 1-SD increase), and 10 were associated with reduced CHD risk (OR=0.74 to 0.84 per 1-SD increase; all FDR<0.1; **Fig. 2a**). Among these 24 metabolites, 21 showed concordant directions of associations with UPF intake and CHD incidence: 12 metabolites positively associated with UPF intake were linked to increased CHD risk and nine metabolites negatively associated with UPF intake were linked to reduced CHD risk.

**Fig. 2.**
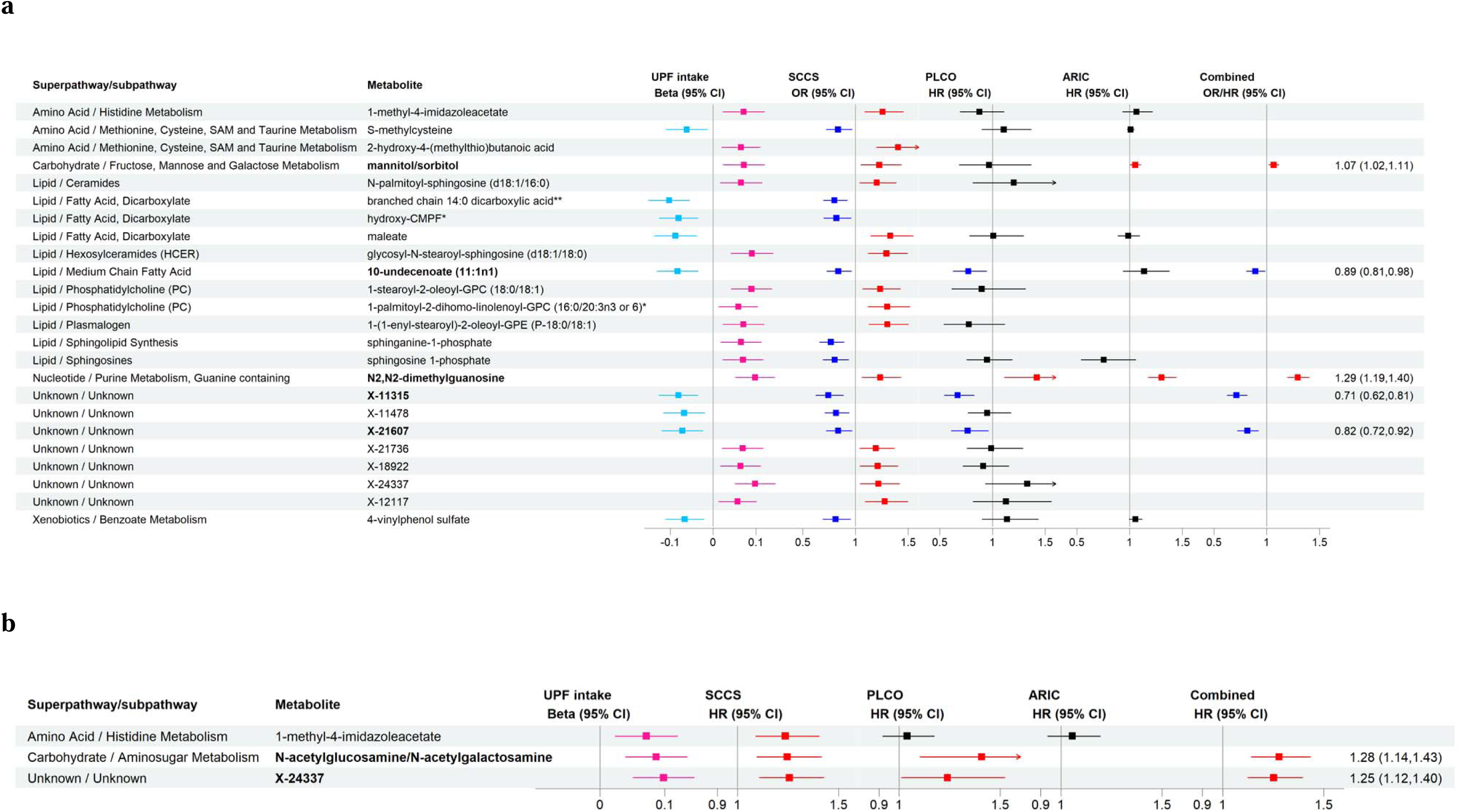
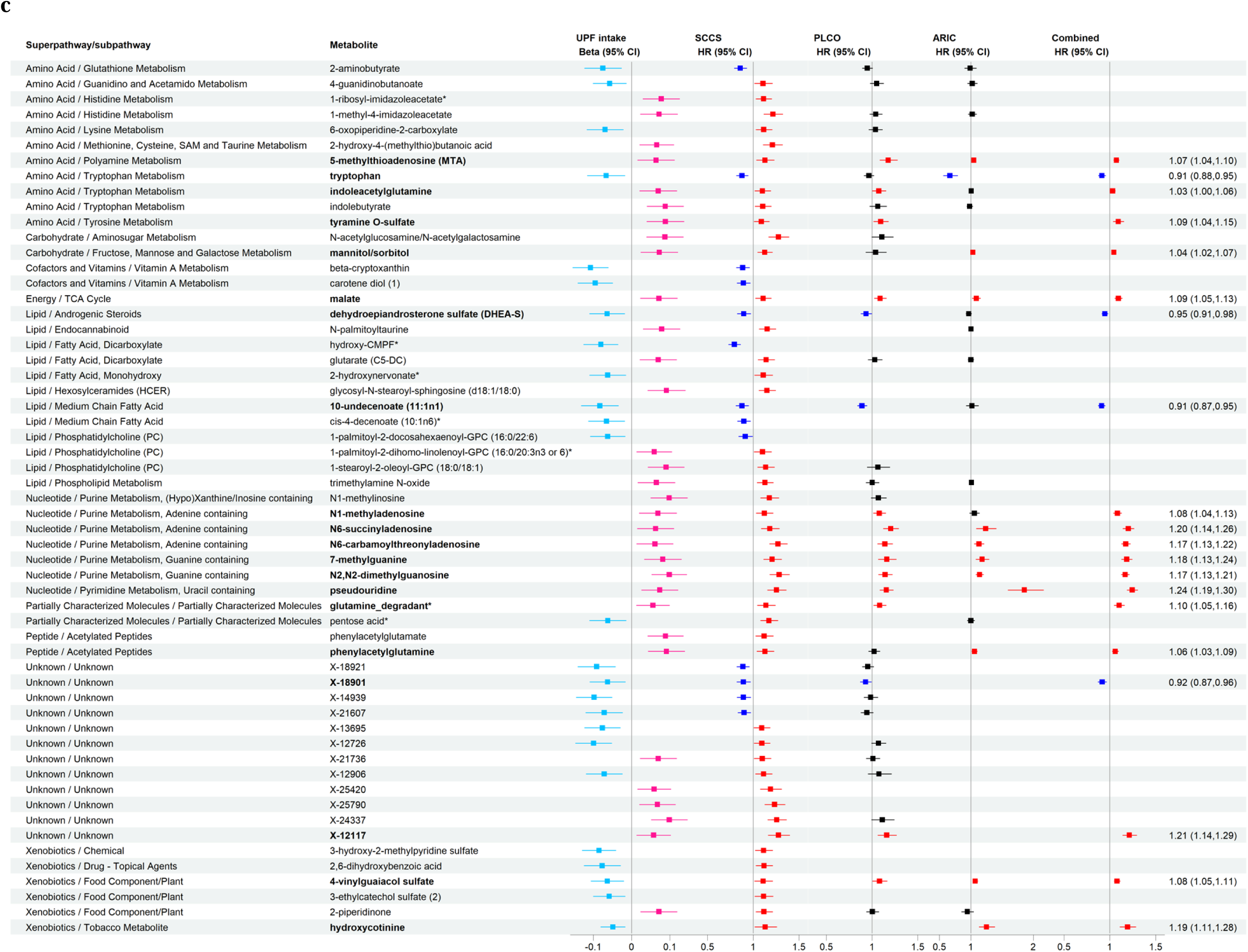
Discovery and Validation of UPF-related metabolites with CHD incidence/mortality, CVD mortality, and total mortality in SCCS, PLCO, and ARIC. a. CHD incidence/mortality. b. CVD mortality. c. Total mortality In the SCCS, UPF-related metabolites showing significant associations with CHD incidence (n=24), CVD mortality (n=3), and total mortality (n=57) at FDR <0.1 were presented and proceeded for validation. A metabolite with a P value < 0.05 and the same direction of association as the SCCS results in either PLCO or ARIC was considered validated. Associations of validated metabolites (in bold) with outcomes in SCCS, PLCO, and ARIC were combined by fixed-effect meta-analysis.

In the PLCO, 87 CHD deaths were documented during an average follow-up of 20 years; in the ARIC study, 577 incident CHD cases were identified during an average follow-up of 24 years. Among those 24 metabolites identified in the SCCS, 18 metabolites were available for validating their associations with CHD incidence or mortality, and 5 metabolites showed significant associations (P<0.05 in either validation cohort; **Fig. 2a**), including N2,N2-dimethylguanosine from nucleotide (combined OR/HR [95%CI] of three studies—SCCS, PLCO, and ARIC: 1.29 [1.19, 1.40] per 1-SD increase), mannitol/sorbitol from carbohydrate (1.07 [1.02, 1.11]), 10-undecenoate from lipid (0.89 [0.81, 0.98]), and two unnamed metabolites (0.71 [0.62, 0.81] for X-11315 and 0.82 [0.72, 0.92] for X-21607).

### UPF-related metabolites with CVD and total mortality

A total of 285, 316, and 744 CVD deaths were documented during mean follow-ups of 13, 20, and 25 years in SCCS, PLCO, and ARIC, respectively. Three UPF-related metabolites were significantly associated with CVD mortality in the SCCS (HR=1.24 to 1.26; all FDR<0.10), two of which were validated in the PLCO: N-acetylglucosamine/N-acetylgalactosamine from amino sugar (combined HR [95%CI]: 1.28 [1.14, 1.43]) and an unnamed metabolite (X-24337; combined HR: 1.25 [1.12, 1.40]) (**Fig. 2b**).

For total mortality, 677, 1,181, and 2,298 deaths were recorded during mean follow-ups of 13, 20, and 25 years in SCCS, PLCO, and ARIC, respectively. In the SCCS, 57 UPF-related metabolites were significantly associated with total mortality (all FDR<0.10; HR=0.80 to 0.91 for 13 metabolites and 1.09 to 1.29 for 44 metabolites; **Fig. 2c**), with 45 showing concordance in the directions of associations with UPF intake and total mortality. Among these 57 metabolites, 40 were available for validation in PLCO or ARIC, and 20 showed significant associations in the same direction as observed in the SCCS (*P*<0.05 in either validation cohort; **Fig. 2c**): the combined HRs ranged from 0.91 to 0.95 for inverse associations and 1.03 to 1.24 for adverse associations. The validated metabolites were mainly from super pathways of amino acids (n=4) and nucleotides (n=6), involved in the metabolism of polyamine, tryptophan, tyrosine, and purine.

### Metabolite signature (MetSig) of UPF intake in SCCS

To capture an overall metabolite profile related to UPF intake, we conducted elastic net regression among the 142 UPF-related metabolites in the SCCS, which selected 84 metabolites to construct the MetSig for UPF intake (**Supplementary Fig. 1**). The MetSig was significantly correlated with the UPF intake among all participants (r= 0.48; P<0.001; **Supplementary Fig. 2**), and the correlation was consistent across participant subgroups with varied sociodemographic backgrounds, lifestyle behaviors, or metabolic disease history (r= 0.42 to 0.53; all P<0.001; **Supplementary Table 2**).

In the CHD case-control study nested within the SCCS, the UPF intake showed a non-significant positive association with CHD incidence (OR [95%CI]: 1.10 [0.95, 1.27] for per 1-SD increase), whereas the MetSig showed a significant and much stronger association (1.46 [1.23, 1.72]; **Fig. 3a**). In terms of mortality outcomes, both UPF intake and the MetSig were significantly associated with CVD mortality (HR=1.16 vs. 1.27) and total mortality (HR=1.12 vs. 1.25), with the MetSig consistently demonstrating stronger associations. In addition, we found that MetSig mediated a substantial proportion of the associations of UPF intake with CHD incidence (68.2%), CVD mortality (59.2%), and total mortality (77.0%; all P<0.001; **Fig. 3b**).

**Fig. 3.**
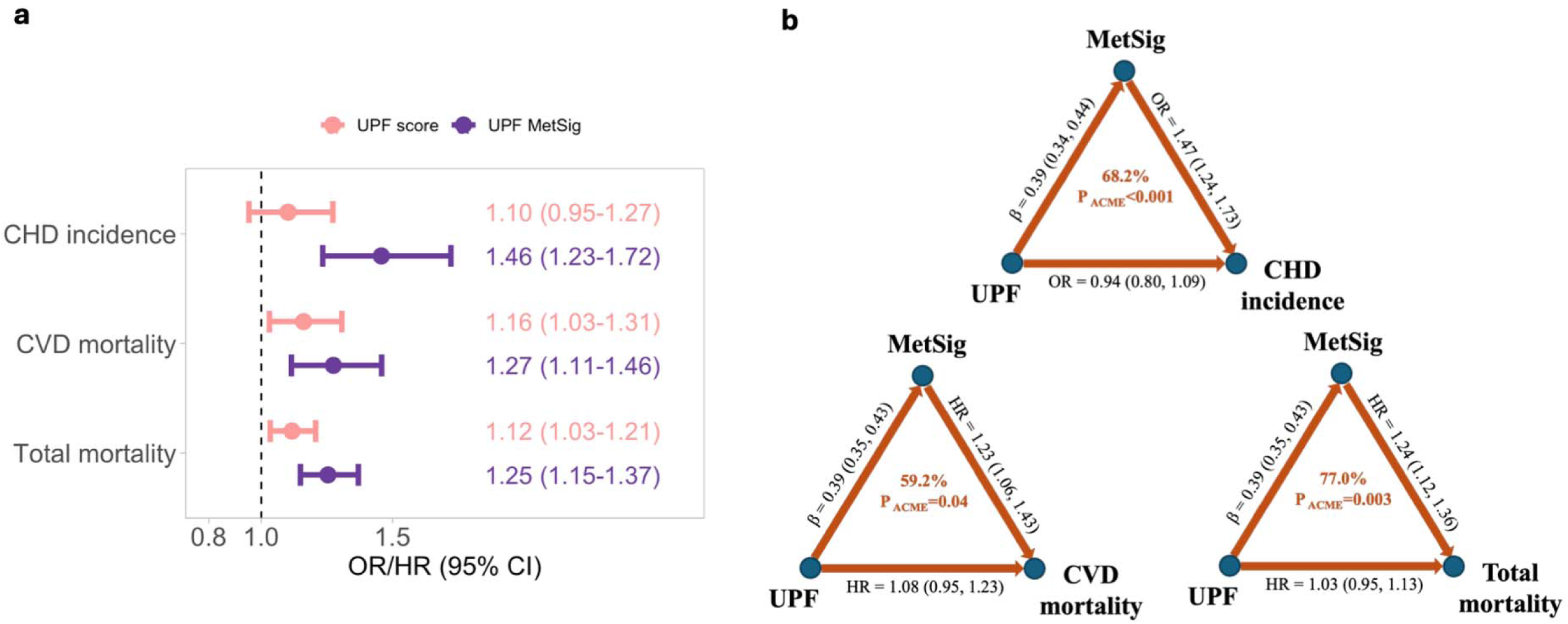
Associations of UPF intake and its MetSig with CHD incidence, CVD mortality, and total mortality in the SCCS. a. Associations of UPF intake and MetSig with CHD incidence, CVD mortality, and total mortality. Conditional logistic regression was used for CHD incidence, and Cox regression was used for CVD and total mortality. Covariates adjusted in the models included technical factors, sociodemographics, lifestyle behaviors, and metabolic health conditions at baseline. b. The mediation effect of MetSig on the associations between UPF intake and outcomes. ACME indicated average causal mediation effect.

Furthermore, adverse associations between the MetSig and health outcomes were consistently observed in participant subgroups regardless of sociodemographics, lifestyle behaviors, obesity status, or metabolic disease history (**Fig. 4**). However, a notably stronger association between MetSig and incident CHD was observed in Black compared to White participants (OR [95%CI]:1.75 [1.37, 2.24] vs. 1.19 [0.95, 1.49]; P for interaction=0.02). Additionally, women and individuals with a history of diabetes or dyslipidemia showed a more pronounced association between the MetSig and total mortality compared to their counterparts (HR [95%CI]: 1.48 [1.25, 1.74] among women, 1.39 [1.19, 1.61] among individuals with diabetes, and 1.40 [1.22, 1.61] among individuals with dyslipidemia; all P for interaction <0.05).

**Fig. 4.**
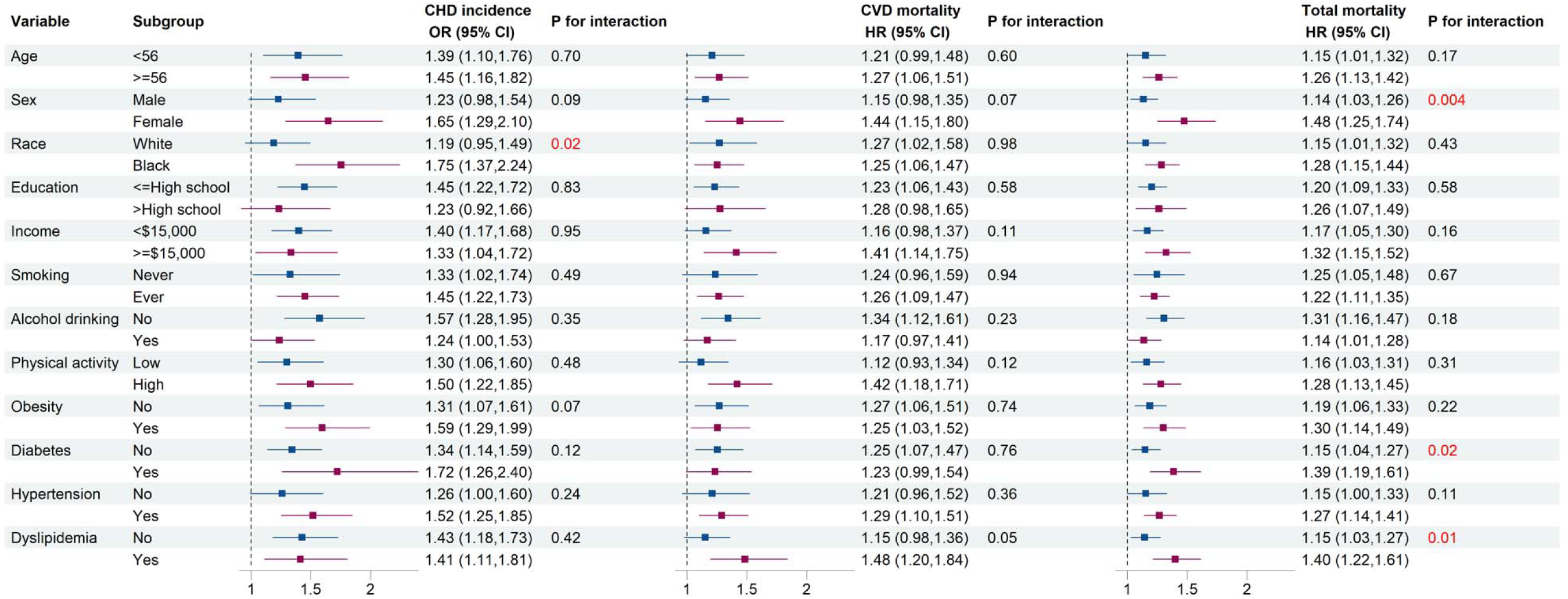
Subgroup analyses for the associations of UPF-MetSig with CHD incidence, CVD mortality, and total mortality in the SCCS. Conditional/regular logistic regression and Cox regression models were used after mutually adjusting for fasting status, batch (for CVD and total mortality only), enrollment age, sex, race, education, income, smoking status, alcohol drinking status, physical activity, sitting hours, daily calories intake, BMI, history of diabetes, hypertension, and hypercholesterolemia at baseline, and family history of CHD (for CHD incidence only).

## Discussion

In this two-stage metabolomics study involving approximately 8,000 Black and White American adults from three large prospective cohort studies, we identified 142 metabolites and validated 43 metabolites associated with UPF intake. We further confirmed significant associations between many of the UPF-related metabolites and adverse health outcomes, including CHD incidence/mortality (n=5), CVD mortality (n=2), and total mortality (n=20). Additionally, in the SCCS, we constructed a metabolite signature for UPF intake comprising 84 metabolites, which consistently demonstrated significant and stronger associations with health outcomes than UPF intake. The metabolite signature mediated 59–77% of the observed UPF-disease associations. Our findings underscore the value of applying metabolomics to elucidate the association and potential biological mechanisms between UPF intake and human health, offering more precise insights than traditional dietary assessments and paving the way for targeted interventions and personalized nutrition strategies to reduce cardiovascular risk and overall mortality.

### UPF-related metabolites

Previous studies have reported a wide and varied range of metabolites being linked to UPF intake, likely reflecting heterogeneity in study populations (e.g., UK, Sweden, or US), UPF assessment methods (e.g., grams/day, servings/day, weight%, or energy%), metabolomic assay platform (e.g., nuclear magnetic resonance [NMR] or liquid chromatography coupled with tandem mass spectrometry [LC-MS]), and statistical analytical approaches (e.g., penalized regression or multivariable linear regression)^12–17^. Several previously reported metabolites were identified and validated in our study, including amino acid metabolites (pipecolate and N-delta-acetylornithine), lipid metabolite (DHA), xenobiotic metabolites (4-vinylphenol sulfate, 3-hydroxyhippurate, catechol sulfate, 4-allylphenol sulfate, and methyl glucopyranoside [alpha + beta]), and nucleotide metabolite (N2, N2-dimethylguanosine)^13–16^. Interestingly, our study noted a negative association between UPF intake and stachydrine (a biomarker of citrus fruits) in both SCCS and PLCO, contrasting with the positive association in the ARIC study^13^, possibly due to the contribution of sweetened fruit drinks to UPF intake and differences in UPF measurement (weight% in SCCS and PLCO vs. serving/day in ARIC). In addition, our study validated other metabolites that decreased with higher UPF intake, including long-chain omega-3 fatty acids DPA and DHA and several polyphenol metabolites (e.g., 3-methoxycatechol sulfate, 3-(3-hydroxyphenyl)propionate sulfate, 3-hydroxypyridine sulfate, ferulic acid 4-sulfate, and quinate)^18^. Conversely, several metabolites increased with higher UPF intake, including anthranilate, indoleacetylglutamine, and 3-indoxyl sulfate from tryptophan metabolism (an essential amino acid abundant in protein-rich foods such as meat, poultry, and cheese; despite tryptophan itself showing an significantly inverse association with UPF in the SCCS) and multiple metabolites from purine (a nitrogen-containing compound essential for DNA and RNA structure and commonly found in red meat, organ meats, and certain seafood).

### UPF-related metabolites and outcomes

Among the UPF-related metabolites, we identified and validated four named metabolites associated with CHD incidence or CVD mortality: mannitol/sorbitol (UPF [+]), N2, N2-dimethylguanosine (UPF[+]), and N-acetylglucosamine/N-acetylgalactosamine (UPF[+]) were all linked to increased risk (combined OR/HR =1.07-1.29), whereas 10-undecenoate (UPF[-]) was linked to decreased risk (combined OR/HR =0.89). Mannitol and sorbitol are sugar alcohols commonly used as low-calorie sweeteners in many UPFs. Higher plasma levels of mannitol/sorbitol have been associated with a 15% increased risk of CHD per 1-SD increment in the Nurses’ Health Study^19^. N2, N2-dimethylguanosine, a byproduct of RNA turnover, reflects oxidative stress and has been linked to incident CHD in other large US cohorts such as the Jackson Heart Study (HR=1.32) and Women’s Health Initiative (HR=1.18)^20^. N-acetylglucosamine and N-acetylgalactosamine are amino sugars involved in glycosylation processes that significantly impact cardiovascular health by modulating inflammation, lipid metabolism, and protein function^21^. Alterations in glycosylation pathways can contribute to CVD pathogenesis. In contrast, 10-undecenoate, a monounsaturated fatty acid, may play a protective role, potentially serving as a biomarker of dairy fat intake^22^, although its cardioprotective mechanisms remain underexplored.

Twenty UPF-related metabolites were also validated for their associations with total mortality, mainly from amino acids (n=4) and nucleotides (n=6). Among the amino acid metabolites, only tryptophan (UPF [-]) was linked to reduced total mortality (combined HR=0.91). As an essential amino acid involved in the synthesis of serotonin and melatonin, tryptophan impacts sleep, mood, and cognitive function. High tryptophan levels have been associated with reduced total mortality in the Malmö Diet and Cancer-Cardiovascular Cohort (HR=0.92)^23^. Conversely, three amino acid metabolites were linked to increased mortality in our study (combined HR=1.03-1.09), including 5-methylthioadenosine (UPF [+]), indoleacetylglutamine (UPF [+]), and tyramine O-sulfate (UPF [+]). Emerging evidence suggests these metabolites may contribute to adverse health outcomes. For example, 5-methylthioadenosine is a byproduct of polyamine metabolism and plays roles in cellular proliferation and apoptosis. Elevated levels of 5-methylthioadenosine have been observed in patients with leukemia, suggesting a potential role in cancer progression^24,25^. Indoleacetylglutamine, a microbial metabolite of tryptophan, has recently been associated with cardiovascular morbidity and mortality in cohorts of the US GeneBank, European LipidCardio, and Qatar Biobank^26,27^. For tyramine O-sulfate, evidence remains limited, underscoring the need for further investigation. Notably, several modified nucleosides involved in purine and pyrimidine metabolism were associated with increased mortality in our study (combined HR=1.08-1.24), including N1-methyladenosine, N6-succinyladenosine, N6-carbamoylthreonyladenosine, 7-methylguanine, N2,N2-dimethylguanosine, and pseudouridine, all of which were positively associated with UPF intake. Their presence in blood or urine reflects cellular RNA turnover, which increases during cell stress, inflammation, oxidative damage, cancer, and aging. Previous studies have consistently reported adverse associations between these metabolites and total mortality^28–32^.

### Metabolite signature of UPF intake

To evaluate the aggregated influence of UPF-associated metabolites on cardiovascular health and mortality, we constructed a MetSig encompassing a broad range of 84 metabolites from seven pathways and demonstrated its significant associations with CHD incidence, CVD mortality, and total mortality. Notably, these associations were stronger for MetSig compared to UPF intake assessed using food frequency questionnaires [FFQ], particularly for CHD incidence (OR [95%CI]: 1.46 [1.23, 1.72] vs. 1.10 [0.95, 1.27]), suggesting that the MetSig captures more information related to levels of food processing than traditional dietary assessment methods (e.g., FFQ). In the Malmö Diet and Cancer cohort, UPF intake was also associated with a modest increase in total mortality (HR [95%CI]: 1.03 [1.02-1.05]), while the metabolite signature of UPF showed a stronger association (1.23[1.06,1.42])^15^. Furthermore, our study found that the MetSig mediated a substantial proportion (59-77%) of the associations between UPF intake and disease outcomes, underscoring its biological relevance. These findings suggest that the metabolite signature may not only reflect dietary intake but also capture intermediate biological pathways through which UPFs influence disease risk. These results highlight the potential of metabolomic signature as an objective, mechanistic biomarker to enhance the accuracy and interpretability of nutritional epidemiology findings.

We observed robust adverse associations between the MetSig and disease outcomes across subpopulations grouped by sociodemographics, lifestyle behaviors, or status of obesity and metabolic diseases. However, the association appeared stronger among females than males, which may reflect differences in metabolism and hormonal regulation^33,34^. Similarly, the stronger association in Black compared to White individuals may be attributed to disparities in diet quality and UPF consumption^35^, a higher burden of comorbidities^36,37^, or barriers to healthcare access, all of which could amplify the adverse metabolic and cardiovascular consequences of UPF intake. Furthermore, the more pronounced associations among individuals with pre-existing metabolic conditions suggest that these individuals may be more vulnerable to the harmful effects of high UPF intake, underscoring the benefit of targeted dietary interventions for high-risk populations. These potential effect modifications warrant further investigation to better understand the role of demographic and clinical factors in shaping UPF– metabolite–disease relations.

### Strengths and limitations

Our study has several notable strengths. It is a large, two-stage metabolomics study involving discovery and validation among ∼8,000 Black and White American adults from three well-established and diverse cohorts (SCCS, PLCO, and ARIC), strengthening the rigor and generalizability of our findings. We identified and validated a broad spectrum of metabolites related to UPF intake and prospectively examined their associations with multiple disease outcomes, providing insights into the potential biological mechanisms underlying the adverse health effects of UPF. Furthermore, we constructed a metabolite signature to capture integrated metabolic profiles related to UPF intake, enhancing the precision to assess UPF-disease relationships beyond self-reported intake. Lastly, all three cohorts collected extensive data on sociodemographic factors, lifestyle behaviors, and medical history, enabling adjustment for potential confounders. However, some limitations should be acknowledged. First, although dietary data were collected using FFQ in all three cohorts, the UPF measurement methods were different (weight% in SCCS and PLCO vs. serving/day in ARIC), which may limit the comparability and validation of findings. Second, the study participant selection was different in three cohorts, with nested case-control designs in the SCCS and PLCO and a cohort design in the ARIC study, potentially introducing heterogeneity in the observed associations. Third, although all metabolomics data were measured using Metabolon’s DiscoveryHD4 platform, some metabolites identified in the SCCS were not available in PLCO or ARIC for validation, probably due to differences in batch processing over time. Fourth, although we have adjusted for a wide range of covariates, residual confounding from unmeasured or poorly measured variables cannot be entirely ruled out.

In conclusion, by leveraging blood metabolomics data from three large prospective cohorts, we identified and validated a set of metabolites across key super pathways (e.g., amino acids, lipids, nucleotides, and xenobiotics) for UPF intake, several of which were linked to CVD morbidity and mortality, as well as total mortality. We also constructed a metabolite signature for UPF intake, which demonstrated strong associations with disease outcomes and explained a significant proportion of the UPF-disease associations. Our findings highlight the potential of metabolomics to provide more objective and biologically grounded insights into the complex relations between UPF and health, supporting its utility in advancing precision nutrition and informing targeted interventions to reduce cardiometabolic risk and improve population health.

## Methods

### Study design and population

This is a two-stage study, including a discovery stage conducted in the SCCS and a validation stage conducted in PLCO and ARIC. The study design and procedures of SCCS, PLCO, and ARIC have been described previously^38–40^. In brief, the SCCS is a prospective cohort study that enrolled 84,735 Americans aged 40–79 years from 12 southeastern states during March 2002 – September 2009^38^. About two-thirds of participants self-identified as Black/African American, and over 50% reported an annual household income of <$15,000. For the current analysis, 1,688 participants were selected from two metabolomic nested case-control studies within the SCCS: one of incident CHD (N=1,023) and one of incident prostate cancer (N=655)^41^. For both studies, the included participants had no history of cancer and provided blood samples at baseline. Participants with missing dietary information or implausible daily energy intake (<600 or >8,000 kcal/day) were excluded. For the CHD case-control study, additional inclusion criteria were: 1) no history of CHD, stroke, heart failure, atherosclerosis, or chronic kidney disease at baseline; 2) no antibiotics use or flu/cold symptoms in the 7 days before blood collection; and 3) eligibility for Centers for Medicare & Medicaid Services (CMS), with ≥ 2 claims after cohort enrollment through December 2016 to facilitate CHD adjudication. CHD cases were 1:1 matched with controls who were free of CHD, heart failure, stroke, or cancer at the time of case diagnosis, matched by race, sex, enrollment age (±2 years), fasting time (±2 hours), and time between sample collection and lab processing (±4 hours).

The PLCO is a randomized, controlled trial that assigned approximately 155,000 participants (50% men and 85% White) to the control arm or the cancer screening intervention arm during 1993–2001^39^. Eligible participants were aged 55–74 years at baseline and had no history of prostate, lung, colorectal, or ovarian cancer. In the intervention arm, participants completed the dietary questionnaire (DQX) at baseline and provided blood samples annually from T0 to T5, except at T3. The present validation analysis focused on 2,725 participants in the intervention arm who were previously selected for four metabolomic nested case-control studies of cancer risk (colorectal: 255 cases and 223 controls; esophageal: 131 cases and 131 controls; breast: 619 cases and 617 controls; prostate: 373 cases and 376 controls)^42–44^. All selected participants were free of cancer at time of blood collection. We further excluded participants who had missing dietary data or implausible daily caloric intake <600 or >8,000 kcal; N=130) or who reported a diagnosis of CHD or stroke at baseline (N=280), leaving 2,315 participants in the final analysis.

The ARIC study is a prospective cohort study that enrolled 15,792 participants (∼73% White) aged 45–64 years from four US communities (Forsyth County, NC; Jackson, MS; suburbs of Minneapolis, MN; Washington County, MD) during 1987 – 1989^40^. The present validation analysis focused on 4,032 participants with blood metabolomics data at baseline^13^. We further excluded participants with missing dietary data or implausible daily caloric intake (<600 or >8,000 kcal; N=2), as well as those who reported a history of CHD at baseline (N=348), leaving 3,682 participants in the final analysis.

All cohorts had institutional review board (IRB) approval from all participating institutions, and the present study was approved by the IRB of Vanderbilt University Medical Center. All participants provided written informed consent.

### Assessment of UPF intake

Usual dietary intake over the past year was collected at baseline using food frequency questionnaires (FFQ) in SCCS (89 items), PLCO (137 items), and ARIC (66 items). Based on the Nova classification system, all FFQ items were categorized into four groups according to the extent and purpose of food processing: 1) Group 1 — un/minimally processed foods, including natural edible parts of plants or animals; 2) Group 2 — processed culinary ingredient, including substances obtained directly from group 1 foods or from nature and used in cooking to prepare meals (e.g., oils, butter, sugar, salt, honey, starch); 3) Group 3 — processed foods, which are products made by adding Group 2 ingredients to Group 1 foods using simple processes of preservation or cooking; and 4) Group 4 — ultra-processed foods, which are industrial formulations made mostly or entirely from substances derived from foods and additives, containing little or no whole foods (e.g., breakfast cereals, reconstituted meat products, carbonated drinks)^1^. In the SCCS, 46 food items were classified as UPF, with 32 items assigned a 100% weight and 14 assigned correction weights ranging from 20% to 70%, based on the likelihood that a given food or food group was ultra-processed in the SCCS population^35^. In the PLCO, 53 food items were classified as UPF, with 40 items assigned a 100% weight and 13 assigned correction weights between 20% – 70% (**Supplementary Table 3**). In the ARIC study, 22 food items were classified as UPF, all assigned a 100% weight^45^. UPF intake was calculated as the percentage of total daily food intake by weight (grams) in the SCCS and PLCO, and as servings/day in the ARIC study.

### Metabolite profiling

Baseline plasma samples of selected SCCS participants were retrieved from the Vanderbilt Molecular Epidemiology Core. Untargeted metabolite profiling was performed by ultra-high-performance liquid chromatography coupled with tandem mass spectrometry (UHPLC-MS/MS) using the Discovery HD4 platform at Metabolon, Inc. (Morrisville, NC, USA) following a standard assay protocol^46^. In brief, plasma samples were extracted with methanol and divided into four aliquots for UHPLC-MS/MS assays in positive and negative ion modes using a combination of reverse phase and HILIC chromatography methods. Metabolites were identified through automated comparison of mass spectra features with the reference library of >4,000 authenticated standard compounds followed by visual inspection for quality control. The peaks were quantified by area under the curve. In total, we identified 1,502 metabolites in selected samples, and 1,110 metabolites detected in ≥ 80% of participants were included in our analysis. Metabolites with missing values were imputed by half of the minimum values in the non-missing samples. Metabolite data were log-transformed and standardized to Z-scores (mean=0 and SD=1).

The metabolite data available in PLCO and ARIC have been published previously^13,42–44^; both conducted untargeted metabolite profiling using UHPLC-MS/MS at Metabolon, the same method as used in the SCCS. In the PLCO, non-fasting blood samples collected at baseline or 1-2 years after baseline were used to measure a total of 2,099 metabolites across the four nested case-control studies. In the ARIC study, fasting blood samples were collected at baseline, and untargeted serum metabolomic profiling was performed in two non-overlapping batches, in 2010 and 2014. Batch 1 consisted of 1,767 Black participants from the Jackson site, and Batch 2 included 1,915 Black and White participants from all four sites. A total of 360 named, non-drug metabolites were available in both batches. Metabolite data in PLCO and ARIC were rescaled to set the median equal to 1. Metabolites with missing values were imputed by the observed minimum values, and imputed metabolites were log-transformed and standardized to Z-scores (mean=0 and SD=1).

### Outcome ascertainment

The outcomes of interest included incident CHD, CVD mortality, and total mortality. In the SCCS — CHD nested case-control study^8^, incident CHD (nonfatal events and CHD deaths) were identified based on International Classification of Diseases (ICD) and Current Procedural Terminology (CPT) codes through CMS claims and National Death Index (NDI). Nonfatal CHD events included acute myocardial infarction (ICD-9 code: 410), coronary revascularization (ICD-9 codes: 36.0-36.3 or 00.66; CPT codes: 92980-92996 or 33510-33536), and other CHD (ICD-9 codes: 411, 413, or 414); CHD deaths were identified by ICD-9 codes 410-414. For CVD mortality (ICD-10 codes I00 I78) and total mortality, vital status, age at death, and underlying cause of death were ascertained in the SCCS via linkage to the National Death Index (NDI) and/or Social Security Administration mortality files up to Dec. 31, 2020.

In the PLCO, all deaths and causes of death were ascertained through Annual Study Update form and periodic linkage to the NDI up to Dec. 31, 2022. The final cause of death was grouped according to ICD-9 codes. The primary endpoints in the validation analysis were CHD mortality (including deaths from ischemic heart disease), CVD mortality (including deaths from ischemic heart disease, cerebrovascular accident, and other circulatory diseases), and total mortality. In the ARIC study, incident CHD (defined as definite or probable myocardial infarction, silent myocardial infarction detected by ECG, coronary revascularization, and definite fatal CHD)^47^, CVD mortality (ICD-9 codes 390 to 459 or ICD 10 codes I00 to I99) and total mortality were ascertained through Dec. 31, 2022.

### Statistical analysis

#### Discovery stage in SCCS

In the SCCS, the associations between UPF intake (per 1-SD increase) and 1,110 individual metabolites were evaluated using linear regression models, adjusting for: 1) technical factors — fasting status (< or ≥8 hours) and batch; 2) sociodemographics — enrollment age (years, continuous), sex (male or female), race (Black or White), education (less than high school, graduated high school, some college, or graduated college or higher), and annual household income (<$15,000, $15,000 – $24,999, $25,000 – $49,999, or ≥ $50,000); 3) lifestyle behaviors — smoking status (never, former, or current), pack years (continuous), number of alcohol drinks per day (continuous), physical activity (metabolic equivalent of task [MET] hours/day, continuous), sitting hours (continuous), and total daily calories (kcal, continuous); and 4) metabolic health conditions — BMI (kg/m^2^, continuous) and history of diabetes (yes or no), hypertension (yes or no), and hypercholesterolemia (yes or no) at baseline. Benjamini and Hochberg false discovery rates (FDR) were calculated, and an FDR <0.1 was considered statistically significant.

Metabolites with FDR<0.1 were further selected by elastic net regression with repeated 10-fold cross-validation to construct the MetSig for UPF intake. The MetSig was calculated as the sum of z scores multiplied by the corresponding regression coefficients. *Pearson* correlation coefficients between UPF score and its MetSig were evaluated among all participants and within subgroups defined by sociodemographics, lifestyle behaviors, and metabolic health conditions.

Subsequently, we evaluated the associations of UPF-related metabolites (FDR<0.1) and MetSig with CHD incidence using conditional logistic regression in the case-control study of CHD, adjusting for enrollment age, education, income, lifestyle behaviors, metabolic health conditions at baseline, and family history of CHD. Cox proportional hazards regression was used to evaluate the associations of UPF-related metabolites and MetSig with CVD mortality and total mortality among all selected SCCS participants, using age as the time scale; entry time was enrollment age, and exit time was age at death or latest follow-up, whichever came first. Covariates in the Cox model included technical factors, sociodemographics, lifestyle behaviors, and metabolic health conditions at baseline. FDRs were calculated among all UPF-related metabolites, and an FDR <0.1 was considered statistically significant.

The causal mediation analysis for the pathway UPF score → MetSig → outcome was conducted using the R packages “mediation” and “mma”, adjusting for covariates as described above. Subgroup analyses for the associations between UPF-MetSig and CHD incidence, CVD mortality, and total mortality were conducted across strata defined by sociodemographic characteristics, lifestyle behaviors, and metabolic health conditions at baseline. Potential effect modification by these stratified variables on the MetSig-outcome associations was examined by adding an interaction term between MetSig and the stratified variable to the regression model.

#### Validation stage in PLCO and ARIC

Metabolites that were significantly associated with UPF intake and subsequently with CHD incidence, CVD mortality, and total mortality at the discovery stage (FDR<0.1) were targeted for validation in PLCO and ARIC. Linear regression was used to assess the associations between UPF intake (per 1-SD increase) and targeted metabolites, and Cox regression was used to assess the associations between targeted metabolites and CHD incidence/mortality, CVD mortality, and total mortality. Covariates included in PLCO were study batch, case-control status, enrollment age, sex, race, education, smoking status, alcohol drinking status, vigorous activity, daily caloric intake, BMI, and history of diabetes and hypertension at baseline; covariates included in ARIC were enrollment age, sex, race-center (for batch 2 only), education, income, smoking status, alcohol consumption, physical activity index, daily caloric intake, BMI, and history of diabetes, hypertension, and lipid-lowering medication use at baseline. In the ARIC study, analyses were conducted separately for the two batches and then meta-analyzed using fixed-effect meta-analysis.

A two-sided *P* value < 0.05 with the same direction of association as the discovery results in either validation cohort (i.e., PLCO or ARIC) was considered validated. Associations of validated metabolites with CHD incidence/mortality, CVD mortality, and total mortality in SCCS, PLCO, and ARIC were meta-analyzed by fixed-effect meta-analysis.

Statistical analyses were conducted using SAS (version 9.4; SAS Institute, Cary, NC, USA) and R (version: 4.3.3).

## Data Availability

All data produced in the present study are available upon reasonable request to the authors.

## Funding Sources

This analysis is supported by R01HL149779 from the National Heart, Lung, and Blood Institute. The Southern Community Cohort Study is funded by U01CA202979 from the National Cancer Institute. Data collection for the Southern Community Cohort Study was performed by the Survey and Biospecimen Shared Resource, which is supported in part by the Vanderbilt-Ingram Cancer Center (P30CA68485). The Atherosclerosis Risk in Communities Study is funded in whole or in part with federal funds from the National Heart, Lung, and Blood Institute, National Institutes of Health, Department of Health and Human Services (75N92022D00001, 75N92022D00002, 75N92022D00003, 75N92022D00004, 75N92022D00005). Metabolomic measurements were funded by the National Human Genome Research Institute (3U01HG004402-02S1). DKG receives research support from the National Heart, Lung, and Blood Institute (R01HL148661). CMR receives research support from the National Heart, Lung, and Blood Institute (R01HL153178). The content is solely the responsibility of the authors and does not necessarily represent the official views of the National Institutes of Health.

## Acknowledgements

The authors thank the staff and participants of the SCCS, PLCO, and ARIC studies for their important contributions and the National Cancer Institute for providing access to PLCO data (Cancer Data Access System [CDAS] project number: PLCO-1820).

## Author contributions

L.W. and D.Y. designed the study. L.W., J.Y., and J.C. analyzed the data. L.W. drafted the manuscript. C.M.R., X.O.S, M.J.S., D.K.G, L.L, W.Z., and D.Y. contributed to the acquisition of data. All authors contributed to the interpretation of results and reviewing and editing the paper and approved the final version of the manuscript. D.Y. is the guarantor of the work and, as such, has full access to all the data in the study and takes responsibility for its integrity and accuracy

## Competing interests

The authors declare no competing interests.

## Notes

### Competing Interest Statement

The authors have declared no competing interest.

### Author Declarations

Ethics committee/IRB of Vanderbilt University Medical Center gave ethical approval for this work.

