## Supplementary file for "Metabolomic Biomarkers and Signature of Ultra-processed Foods Intake and Cardiovascular Morbidity and Mortality"

**Supplementary Table 1. Baseline characteristics of validation population in PLCO and ARIC**

| **Baseline characteristics** | **PLCO  (N=2,315)** | **ARIC**  **(N=3,682)** |
| --- | --- | --- |
| **UPF intake (% or servings/day)^1^** | 22.7 (9.1) | 5.6 (3.0) |
| **Enrollment age (years)** | 63.0 (5.1) | 53.4 (5.7) |
| **Sex** |  |  |
| Male | 1033 (44.6%) | 1440 (39.1%) |
| Female | 1282 (55.4%) | 2242 (60.9%) |
| **Race** |  |  |
| White | 2118 (91.5%) | 1361 (37.0%) |
| Black | 91 (3.9%) | 2321 (63.0%) |
| Others | 106 (4.6%) | / |
| **Educational attainment** |  |  |
| Less than high school | 163 (7.0%) | 1199 (32.56%) |
| High school or vocational school | 831 (35.9%) | 1245 (33.81%) |
| Some college or more | 1321 (57.1%) | 1238 (33.62%) |
| **Smoking status** |  |  |
| Never | 1149 (49.6%) | 1684 (45.74%) |
| Former | 996 (43.0%) | 992 (26.94%) |
| Current | 170 (7.3%) | 1006 (27.32%) |
| **Alcohol drinking^2^** |  |  |
| No | 469 (20.3%) | 1362 (37.0%) |
| Yes | 1846 (79.7%) | 2300 (62.5%) |
| g/week | / | 36.38 (95.5) |
| **Vigorous activities (hours/week)** | 2.19 (1.83) | / |
| **Physical activity index^3^** | / | 2.26 (0.75) |
| **Total energy intake (kcal/day)** | 2050 (841) | 1620 (622) |
| **BMI (kg/m^2^)** | 27.5 (4.8) | 28.8 (5.9) |
| **Diabetes** | 138 (6.0%) | 473 (12.85%) |
| **Hypertension** | 718 (31.0%) | 1619 (43.97%) |
| **Lipid-lowering medication use** | / | 68 (1.9%) |

*Mean ± standard deviation for continuous variables and number (percentage) for categorical variables.

^1^ UPF intake was estimated as the percentage of total daily foods intake by weight (grams) in PLCO and as servings/day in ARIC.

^2^ Alcohol drinking status was assessed as yes/no in PLCO and as current/former/never in ARIC. Current and former drinkers in ARIC were grouped as ‘Yes’.

^3^ Physical activity index was calculated based on leisure-time sport and exercise, accounting for intensity, duration, and frequency; 1 indicates the lowest, and 5 indicates the highest.

**Supplementary Figure 1. Metabolites selected in developing UPF-MetSig in SCCS**

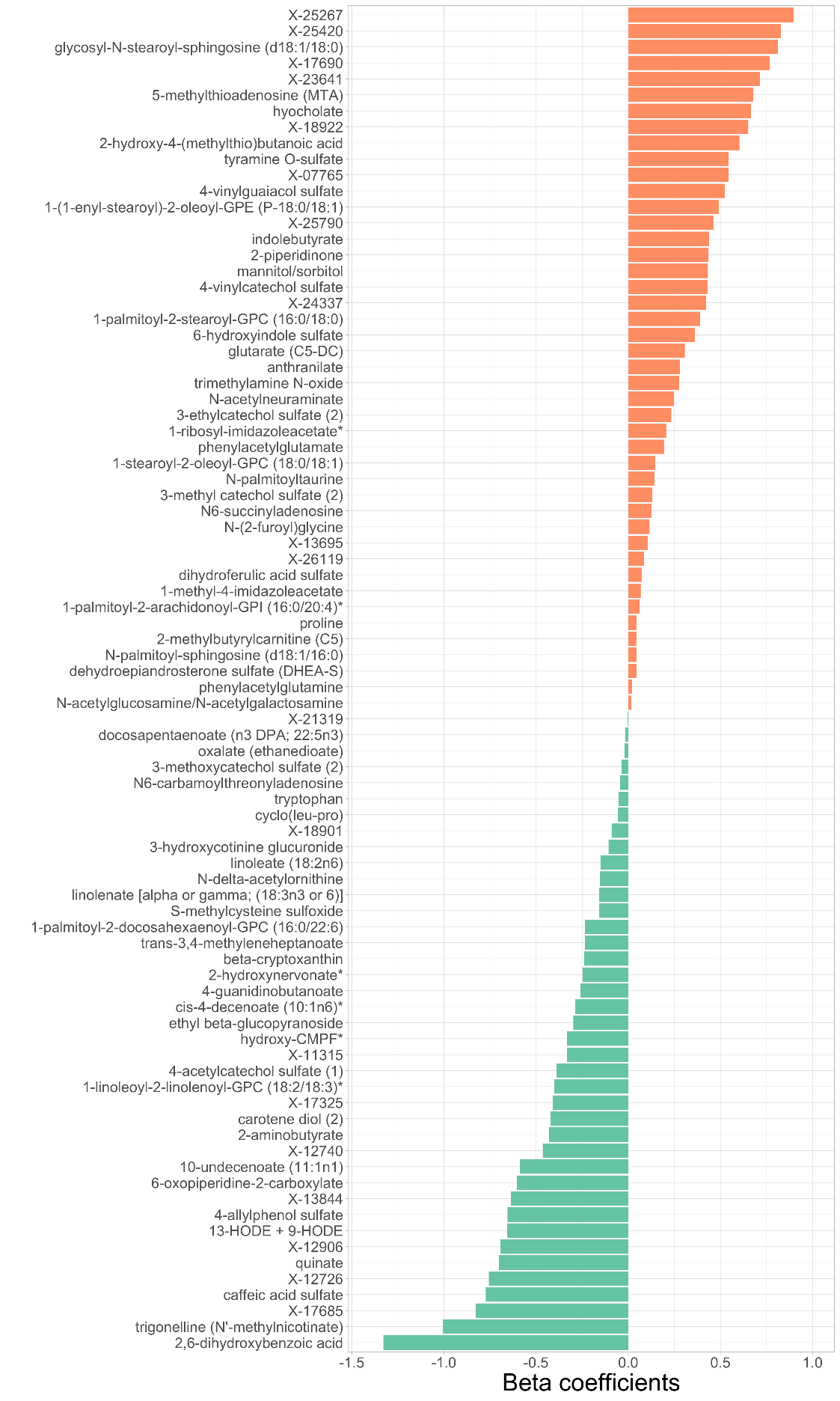

**Supplementary Figure 2. Correlation between UPF intake and its MetSig in SCCS**

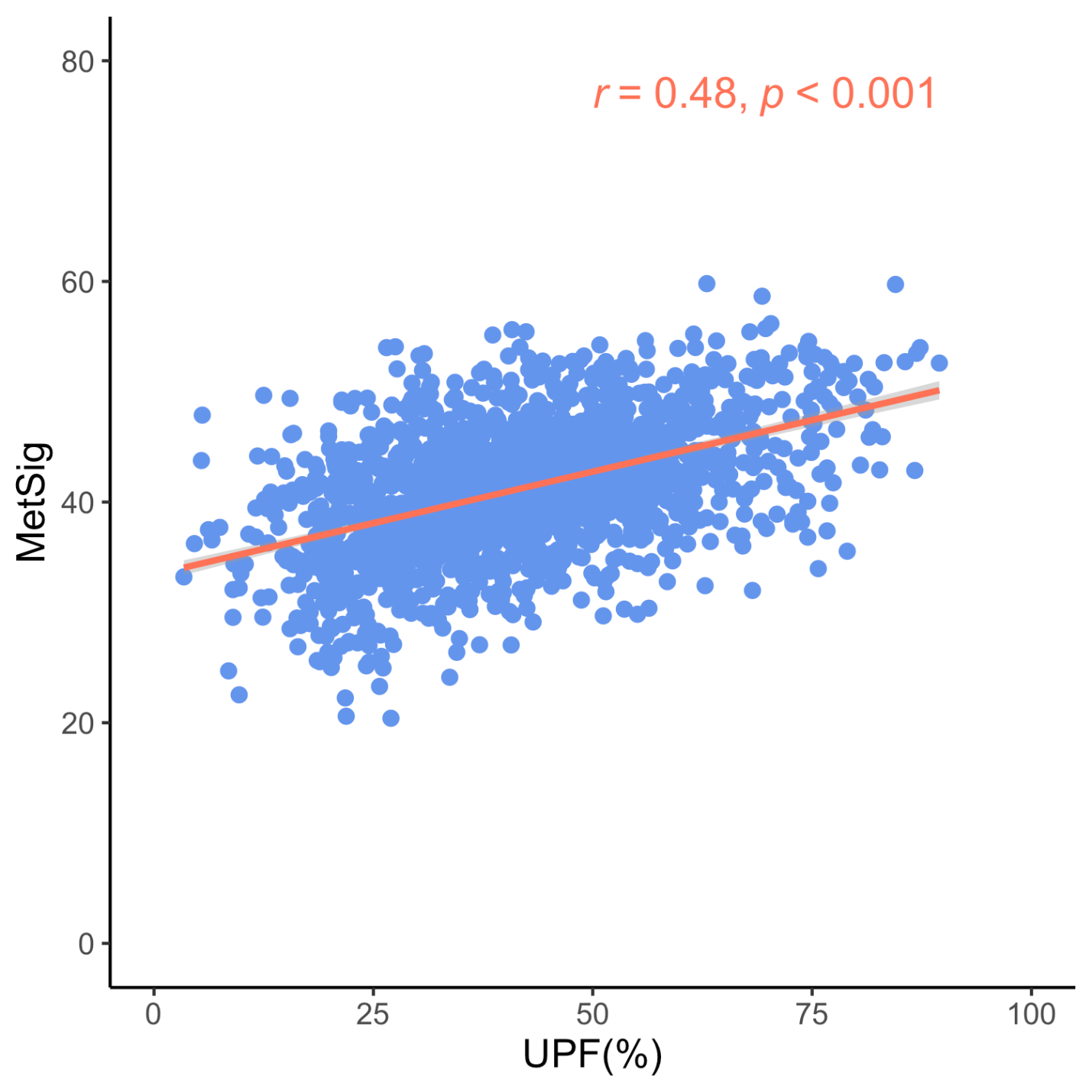

**Supplementary Table 2. Correlations between UPF intake and its MetSig among subgroups in SCCS**

| **Variable** | **Subgroup** | **UPF and MetSig** | |
| --- | --- | --- | --- |
|  |  | ***r*** | ***p*** |
| **Age** | <56 | 0.43 | 5.98E-39 |
|  | ≥56 | 0.48 | 3.88E-51 |
| **Sex** | Male | 0.44 | 5.76E-56 |
|  | Female | 0.53 | 1.13E-38 |
| **Race** | White | 0.53 | 3.29E-50 |
|  | Black | 0.42 | 4.99E-45 |
| **Education** | High school or below | 0.46 | 1.93E-64 |
|  | More than high school | 0.51 | 3.08E-32 |
| **Income** | <$15,000 | 0.47 | 1.89E-56 |
|  | ≥$15,000 | 0.48 | 1.47E-40 |
| **Smoking** | never | 0.48 | 2.44E-28 |
|  | ever | 0.48 | 6.30E-70 |
| **Alcohol drinking** | No | 0.47 | 2.64E-43 |
|  | Yes | 0.49 | 1.83E-55 |
| **Physical activity** | Low | 0.45 | 2.09E-43 |
|  | High | 0.51 | 2.30E-56 |
| **Obesity** | No | 0.49 | 1.16E-60 |
|  | Yes | 0.47 | 7.37E-39 |
| **Diabetes** | No | 0.49 | 3.99E-77 |
|  | Yes | 0.45 | 1.12E-21 |
| **Hypertension** | No | 0.48 | 2.74E-40 |
|  | Yes | 0.48 | 7.02E-59 |
| **Hypercholesterolemia** | No | 0.44 | 3.40E-53 |
|  | Yes | 0.53 | 2.18E-45 |

**Supplementary Table 3. Ultra-processed foods in PLCO**

| **Food** | **Weight** |
| --- | --- |
| Bacon (g/day) | 100% |
| Beef Roast (g/day) | 50% |
| Biscuits and Muffins (g/day) | 100% |
| Corn Bread/Muffins/Tortillas (g/day) | 100% |
| Dark Bread (Wheat,Rye,Pumpernickel) (g/day) | 100% |
| White Bread (g/day) | 100% |
| Cakes (g/day) | 100% |
| Candy, Chocolate (g/day) | 100% |
| Candy, Not Chocolate (g/day) | 100% |
| Hot Breakfast Cereals (g/day) | 50% |
| Ready-to-Eat Cereal, High-Fiber (g/day) | 100% |
| Ready-to-Eat Cereal, Highly Fortified (g/day) | 100% |
| Ready-to-Eat Cereal, Other (g/day) | 100% |
| Ready-to-Eat Cereal, Good Fiber (g/day) | 100% |
| Potato/Corn Chips and Popcorn (g/day) | 100% |
| Coffee (g/day) | 20% |
| Cold Cuts (g/day) | 100% |
| Cookies and Brownies (g/day) | 100% |
| Crackers (g/day) | 100% |
| Sour Cream (g/day) | 100% |
| Sweet Cream (g/day) | 100% |
| Donuts, Sweet Rolls, and Coffee Cake (g/day) | 100% |
| Margarine, Butter, and Oil on Vegetables and Potatoes (g/day) | 33% |
| Fish Excluding Shellfish - Fried (g/day) | 100% |
| Fruit Drinks (g/day) | 100% |
| Gravies Made with Meat (g/day) | 100% |
| Ham (g/day) | 100% |
| Beef Burger (g/day) | 100% |
| Hot Dogs (g/day) | 100% |
| Regular Ice Cream (g/day) | 100% |
| Ketchup and Chili/Taco Sauce (g/day) | 100% |
| Alcoholic Beverages - Liquor and Mixed Drinks (g/day) | 100% |
| Liver and Liverwurst (g/day) | 50% |
| Margarine (g/day) | 100% |
| Meatloaf, Burritos, and Tacos (g/day) | 50% |
| Meat Component Only - Beef Stew (g/day) | 50% |
| Pancakes and Waffles (g/day) | 100% |
| Peanuts and Peanut Butter (g/day) | 35% |
| Other Pies (g/day) | 100% |
| Pumpkin/Sweet Potato Pie (g/day) | 100% |
| Pizza (g/day) | 100% |
| Potatoes - Fried (g/day) | 100% |
| Salad Dressing and Mayonnaise (g/day) | 100% |
| Sausage (g/day) | 100% |
| Soft Drinks and Soda (g/day) | 100% |
| Other Soups (g/day) | 70% |
| Vegetable/Tomato Soup (g/day) | 70% |
| Beef Stew/Pot Pie (g/day) | 50% |
| Beef Steak (g/day) | 50% |
| Tofu and Soybeans (g/day) | 50% |
| Tomato/Spaghetti Sauce (g/day) | 100% |
| White/Cheese Sauce (g/day) | 100% |
| Frozen Yogurt and Ice Milk (g/day) | 100% |
